# Platelet Activation by Vaccine-Induced Immune Thrombotic Thrombocytopenia (VITT) Patient Serum is Blocked by COX, P2Y_12_ and Kinase Inhibitors

**DOI:** 10.1101/2021.04.24.21255655

**Authors:** Christopher W. Smith, Caroline Kardeby, Ying Di, Gillian C. Lowe, William A. Lester, Steve P. Watson, Phillip L.R. Nicolson

## Abstract

**Background:** The novel coronavirus SARS-CoV-2 has caused a global pandemic. Vaccines are an important part of the response. Although rare, unusual thrombotic events and thrombocytopenia in recipients 4–16 days after vaccination with the AstraZeneca AZD1222 have been reported. This syndrome of vaccine-induced immune thrombotic thrombocytopenia (VITT) clinically resembles heparin induced thrombocytopenia (HIT), which is caused by platelet activating antibodies against platelet factor 4 (PF4). Here, we investigate the effect of serum from patients with VITT on platelet activation, and assess the ability of clinically available therapeutics to prevent platelet activation.

**Methods:** Aggregation responses of healthy donor washed platelets were assessed in response to serum from patients with VITT pre- and post-intravenous immunoglobulin (IVIg) treatment and in the presence of anti-FcγRIIA blocking IV.3 F(ab), anti-platelet drugs and kinase inhibitors.

**Findings:** Four patients (21 - 48 years old) presented with thrombosis (three patients: cerebral venous sinus thrombosis, one patient: ischemic stroke) and thrombocytopenia 10-14 days after AZD1222 vaccination. All patients tested positive for anti-PF4 antibody despite no prior heparin exposure. Serum from patients with VITT, but not healthy donor controls, induced platelet aggregation, which was abrogated following IVIg treatment. Aggregation to patient sera was blocked by IV.3 F(ab) which targets FcγRIIA, and inhibitors of Src, Syk and Btk kinases downstream of the receptor. Anti-platelet therapies indomethacin and ticagrelor also blocked aggregation.

**Interpretation:** In conclusion, serum from patients with VITT activates platelets via the FcγRIIA, which can be blocked *in vitro* by anti-platelet therapies suggesting possible new therapeutic interventions for this rare syndrome.

**Funding:** This work was supported by an Accelerator Grant (AA/18/2/34218) from the British Heart Foundation (BHF).

**Key points:**

1. Serum from patients with VITT activates platelets via the FcγRIIA.
2. Platelet activation by serum from patients with VITT can be blocked by COX, P2Y_12_, Src, Syk and Btk inhibition.

## Introduction

The respiratory disease COVID-19, caused by the novel coronavirus, severe acute respiratory syndrome coronavirus 2 (SARS-CoV-2), has caused a global pandemic and millions of deaths. To combat this, several vaccines have been developed.^1–3^ The Oxford-AstraZeneca chimpanzee adenovirus vectored ChAdOx1 nCoV-19 (AZD1222) vaccine has been used in an aggressive vaccination program in the UK and across the European Economic Area due to its low cost and ease of storage (∼20 million people vaccinated as of 08/04/2021).^4^

However, since late February 2021 there have been a growing number reports of unusual thrombotic events with accompanying thrombocytopenia, disseminated intravascular coagulation (DIC) and bleeding with high mortality 4-16 days post vaccination with AZD1222 in young and healthy individuals leading to concerns this is a vaccine-induced condition.^5,6^ This has been termed vaccine-induced immune thrombotic thrombocytopenia (VITT) and has an approximate incidence of 1 in 250,000.^5,7^

VITT has a clinical resemblance to autoimmune heparin induced thrombocytopenia (HIT), with several patients testing positive for anti-platelet factor 4 (PF4) antibodies.^5,8^ In HIT, IgG-containing immune complexes bind and cross-link the platelet surface receptor FcγRIIA (CD32a), a low affinity Fc receptor (FcR) that binds immune complexes with high avidity, and initiate platelet activation.^8^ Autoimmune HIT, despite the name, can rarely occur independently of heparin and result in persistent severe thrombocytopenia together with DIC and microvascular thrombosis.^8^

In this study, we investigated the effect of serum from patients with VITT on platelet aggregation. Serum from patients with VITT induced aggregation of healthy donor platelets *in vitro*. This was abolished following treatment of patients with intravenous immunoglobulin (IVIg) or, *in vitro*, following heat inactivation of patient serum, in the presence of an anti-FcγRIIA monoclonal antibody blocking F(ab), or inhibitors of FcγRIIA downstream signalling kinases and platelet feedback agonists.

## Methods

### Patients and Ethical approval

Patients presenting with thrombosis and thrombocytopenia, occurring after AZD1222 vaccination were recruited. Collection of blood from these patients (including those lacking capacity) was approved under research ethics granted to the University of Birmingham Human Biomaterial Resource Centre (North West – Haydock Research Ethics Committee, reference: 15/NW/0079, amendment 3, 19/11/2018). Ethical approval for collecting blood from healthy volunteers was granted by Birmingham University Internal Ethical Review Committee (reference: ERN_11-0175, assessed 22/3/2021).

### Antibodies and reagents

Mouse monoclonal IgG2b antibody against human CD32 (IV.3) was purified from hybridoma cells supernatant, and IV.3 F(ab) fragment made using Pierce Fab Preparation kit (Thermo Fisher Scientific). Eptifibatide was from GSK. Ibrutinib (PCI-32765), R406 (free base), entospletinib (GS-9973) were from Selleckchem. Rilzabrutinib was provided by Principia BioPharma. All other reagents were purchased from Sigma-Aldrich.

### Serum preparation

Patient and healthy donor serum was collected following centrifugation (2000 × g, 10 minutes, room temperature [RT]) of clotted whole blood. Patient sera was collected before and after treatment with dexamethasone and IVIg (see Table 1).

**Table 1.**
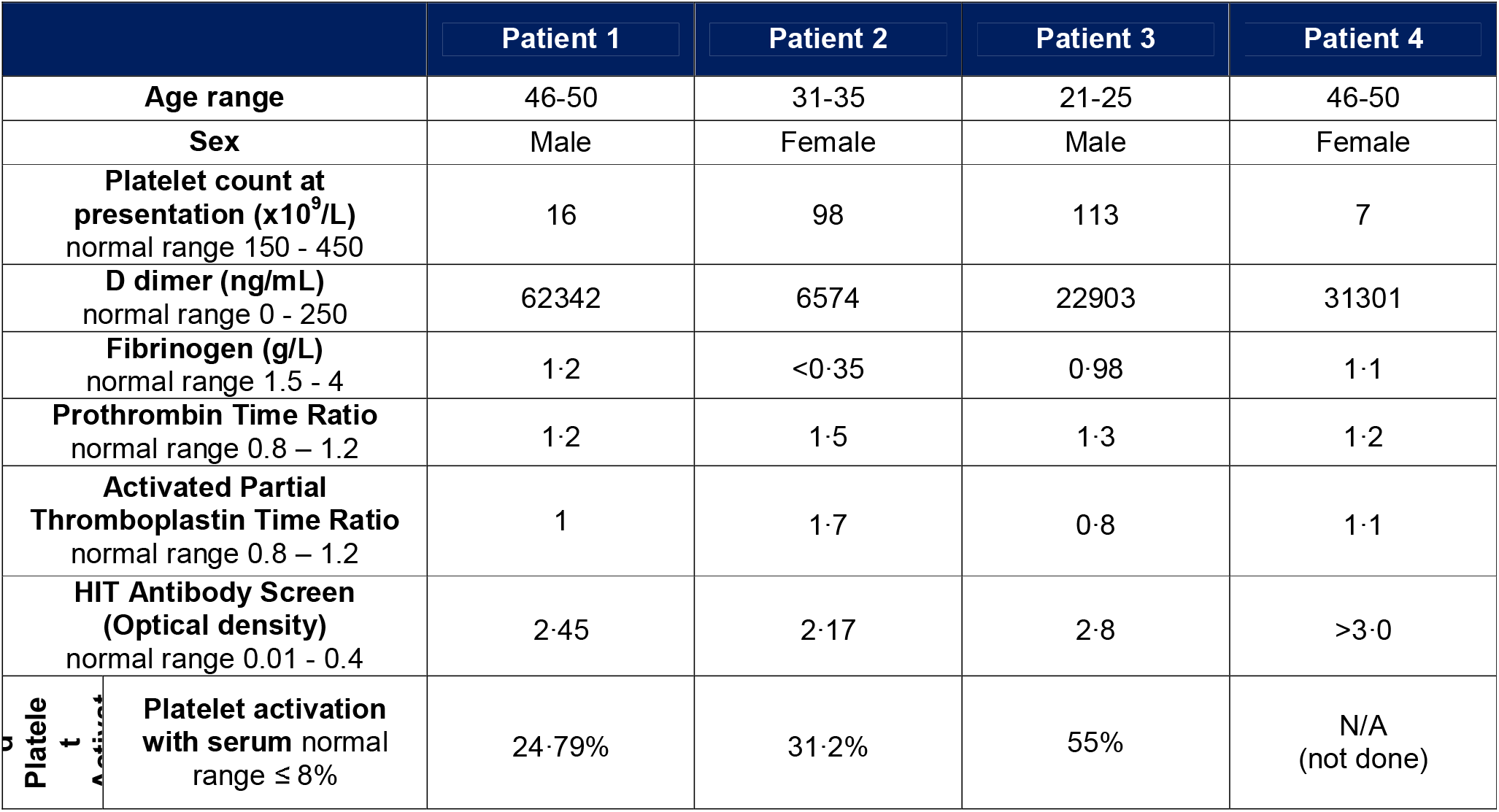
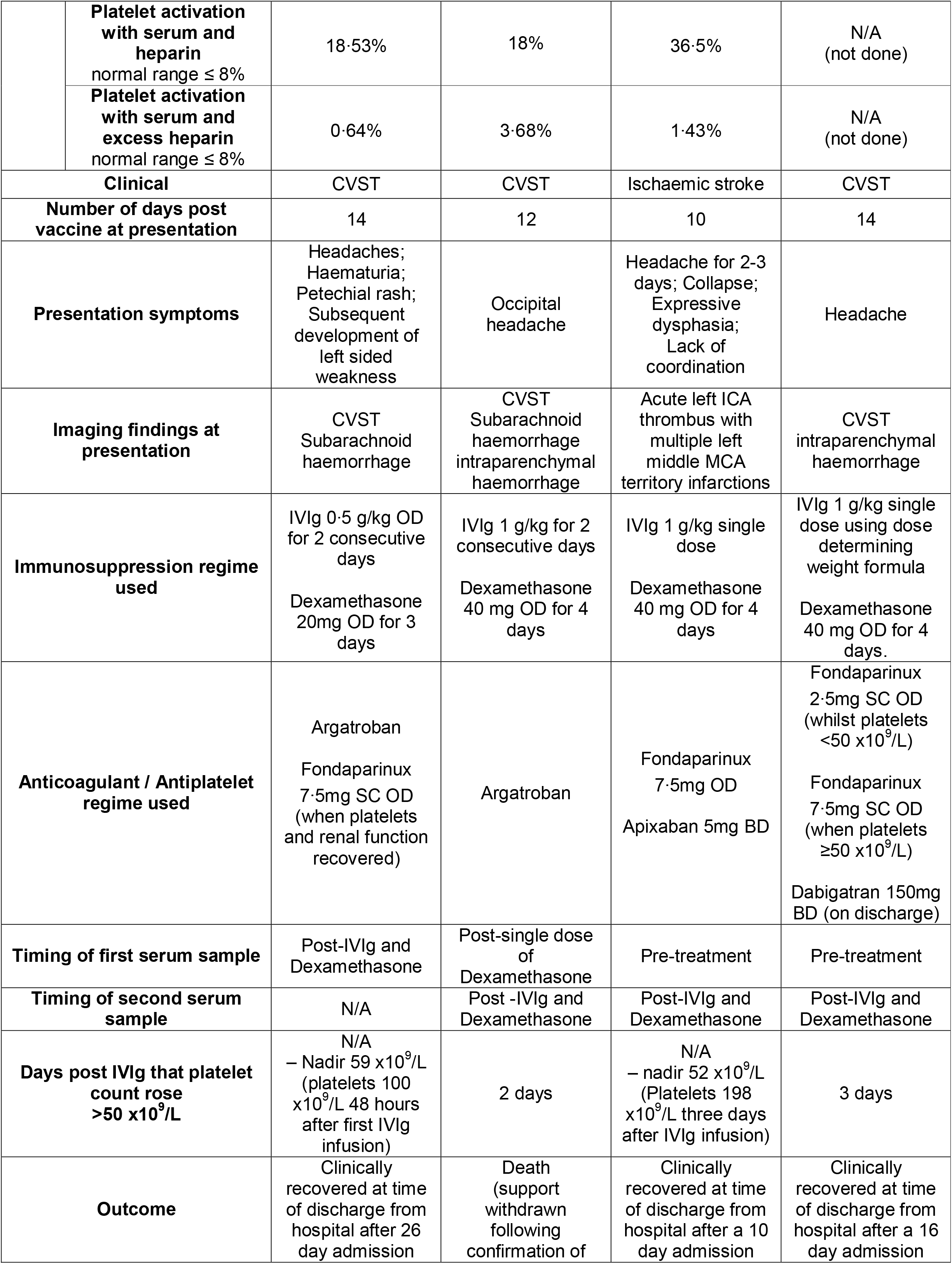
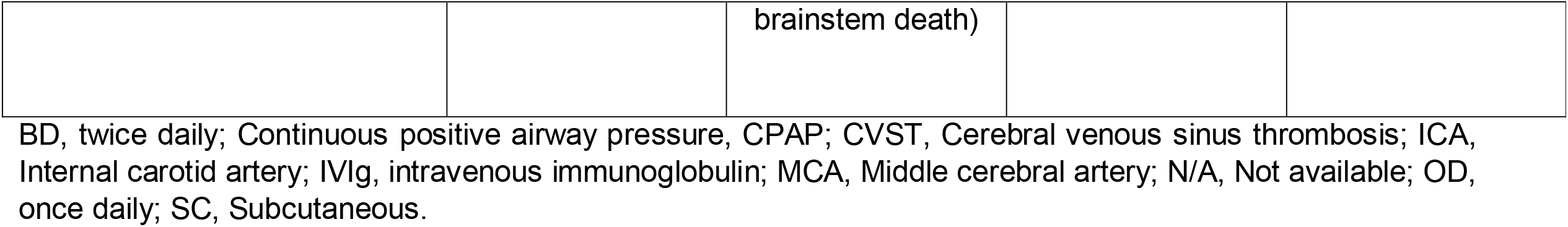
Summary of Patients with Vaccine Induced Thrombotic Thrombocytopenia (VITT).

### Human platelet preparation

Preparation of washed platelets from citrated whole blood has already been described.^9^ Briefly; citrated blood was taken from healthy, drug-free volunteers and mixed (1:10, v/v) with acid citrate dextrose and centrifuged (200 × g, 20 minutes, RT) to produce platelet rich plasma. Platelet rich plasma was then centrifuged (1000 × g, 10 minutes, RT) in the presence of 0.2 μg/ml prostacyclin. The platelet pellet was resuspended in modified-Tyrode’s-HEPES buffer, acid citrate dextrose and 0.2 μg/ml prostacyclin and centrifuged (1000 × g, 10 minutes, RT). Platelet pellet was resuspended in modified-Tyrode’s-HEPES buffer to a concentration of 2×10^8^/ml and allowed to rest for 30 minutes prior to use.

### Light transmission aggregometry (LTA)

Aggregation was measured in washed platelets (2×10^8^/ml) under stirring conditions (1200 rpm) at 37°C using a light transmission aggregometer (Model 700, ChronoLog) for 20 minutes following stimulation with serum (1:15, v/v). Washed platelets were pre-incubated with IV.3 F(ab) for 5 minutes or with inhibitors for 10 minutes prior to stimulation with serum. Tyrode’s or dimethyl sulfoxide (DMSO) was used as vehicle.

### Statistical analysis

All data are presented as mean ± standard error of the mean (SEM), p<0.05 was considered statistically significant. Statistical analysis was performed in GraphPad Prism 9 (GraphPad Software Inc.) using one or two-way ANOVA with Dunnett corrections for multiple comparisons.

## Results

### Patients with VITT demonstrate thrombosis, thrombocytopenia, elevated D-dimer levels and anti-PF4 antibodies on ELISA testing

The presentation, investigation results, treatment and outcomes of four patients with VITT are summarized in Table 1. All patients were Caucasian and under the age of 50, and had not previously had symptomatic COVID-19. All patients presented with headaches, and one also had expressive dysphasia, 10-14 days following dosing with AZD1222. At presentation clinical investigation revealed that all patients were thrombocytopenic (range: 7-113 ×10^9^ platelets/L), with massively elevated D-dimer and low fibrinogen levels. Despite no prior exposure to heparin, HIT screening with the anti-PF4 IgG Immucor assay showed strong reactivity in all patients, with Heparin Induced Platelet Activation (HIPA) assays in the three patients tested showing platelet activation to patient serum that was reduced by low and blocked by high concentrations of heparin. These results are similar to reports of other patients with VITT.^5,6^ Cross-sectional brain imaging verified the presence of cerebral venous sinus thrombosis (CVST) and intracerebral haemorrhage in three patients, and ischemic stroke caused by internal carotid artery thrombus in one patient.

All patients received intravenous immunoglobulin (IVIg) and the steroid dexamethasone, which is recommended by the British Society of Haematology guidelines for VITT^10^, and all had rapid improvements in their platelet counts over 1-3 days. Of note, IVIg has also been shown to rapidly inhibit HIT antibody induced platelet activation.^11^ Patients also received non-heparin anticoagulation, and two patients required intensive care unit support. At the time of writing, three patients had recovered and been discharged from hospital and one patient died in the intensive care unit.

### Serum from patients with VITT induces platelet aggregation

Serum was collected from healthy donors and patients with VITT. Patient 1 had serum collected after IVIg had been administered. Patients 2, 3 and 4 had serum collected both before and after IVIg administration. Patient 2 had received dexamethasone prior to their first serum collection. To investigate the effect on platelet activation, these sera were added to washed platelets and aggregation measured (Figure 1A). Serum from patients with VITT triggered platelet aggregation to variable degrees depending on the platelet donor, which was abolished in post-IVIg treatment sera. Sera from four healthy donor controls did not cause platelet aggregation. Three repeats were performed for each serum sample on platelets from healthy donors known to respond.

**Figure 1.**
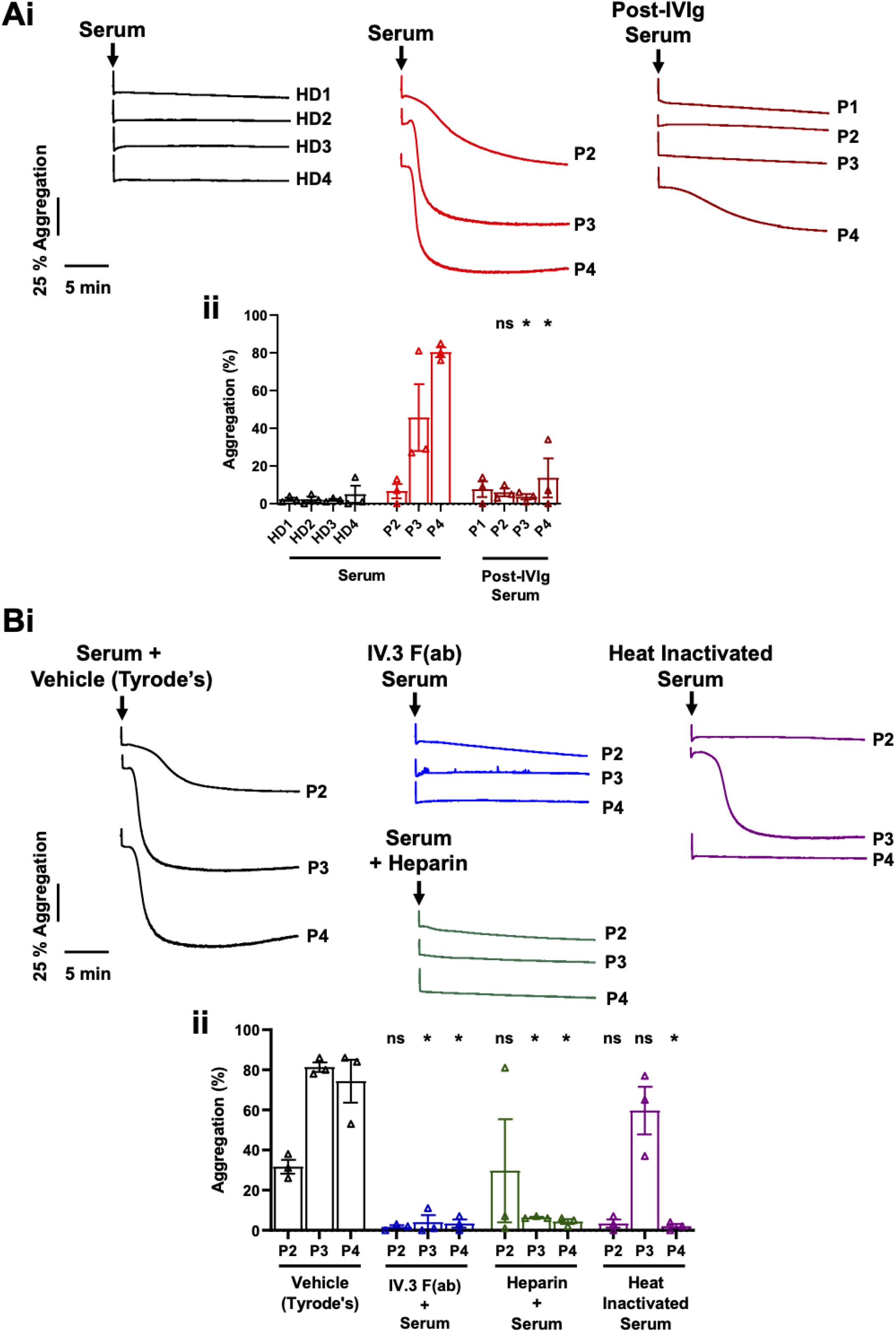
Serum from patients with VITT induces platelet aggregation via the FcγRIIA. Washed platelets (2×10^8^/ml) were stimulated with serum (1:15, v/v) from **(A)** healthy donors (HD) or patients with VITT (P) pre- and post-IVIg treatment, or **(B)** in the presence of 10 μg/ml IV.3 F(ab), low concentration heparin (0.2 U/ml) or following heat inactivation (56°C, 45 minutes) and aggregation measured. **(i)** Representative aggregation traces and **(ii)** quantification of maximum aggregation. Mean ± SEM, n=3. Statistical analysis was by two-way ANOVA with Dunnett multiple comparisons, * p<0.05, ns: non-significant.

Addition of the integrin αIIbβ3 inhibitor eptifibatide (9 μM) inhibited the response to patient sera, confirming that this was aggregation rather than agglutination (data not shown).

### Platelet aggregation to serum from patients with VITT is abolished by FcγRIIA blockade and heat inactivation

Platelet activation in HIT is caused by antibody mediated clustering of FcγRIIA.^8^ To determine if a similar mechanism was involved in VITT we used an anti-FcγRIIA blocking IV.3 F(ab). Platelet activation by patient sera was abolished in the presence of IV.3 F(ab) demonstrating platelet activation in VITT is mediated via FcγRIIA.

The involvement of PF4 and heparin in HIT led us to evaluate their effects in conjunction with patient sera. Both PF4 and low concentrations of heparin have been shown to enhance platelet responses in HIT assays, whereas heparin at high concentrations inhibits any response.^12–14^ No enhancement in the partial aggregation observed to patient 2 serum was observed in the presence of 10 μg/ml PF4 (data not shown). Both low and high heparin concentrations (0.2 U/ml and 100 U/ml, respectively) prevented serum induced aggregation, although a delayed rather than blocked aggregation was observed to low concentrations in a single patient repeat (Figure 1B and data not shown). Three patients (patients 1, 2 and 4) were being given non-heparin anticoagulation with argatroban or fondaparinux at the time of blood collection.

To exclude platelet activation from other sources (such as thrombin and complement) in sera, heat inactivation (56°C, 45 minutes) of the three patient sera that caused activation was used.^15^ Heat inactivation of patient sera blocked aggregation in two patients and had no effect in another patient (Figure 1b).

### Inhibition of cyclooxygenase, P2Y_12_, Src, Syk and Btk blocks platelet aggregation induced by VITT patient serum

Platelet activation by FcγRIIA occurs via Src, Syk and Btk kinases.^16^ We tested if clinically available anti-platelet therapies and inhibitors of these kinases could prevent platelet aggregation to patient sera. The Src inhibitor dasatinib, Syk inhibitor entospletinib and the Btk inhibitors ibrutinib and rilzabrutinib all prevented aggregation to patient sera (Figure 2). R406, the active metabolite of the Syk inhibitor fostamatinib, failed to prevent aggregation. The COX inhibitor indomethacin, which has the same mechanism of action as aspirin, and the P2Y_12_ inhibitor ticagrelor also blocked aggregation. All inhibitors were used at a concentration which fully inhibited aggregation to 3 μg/ml collagen (results not shown).

**Figure 2.**
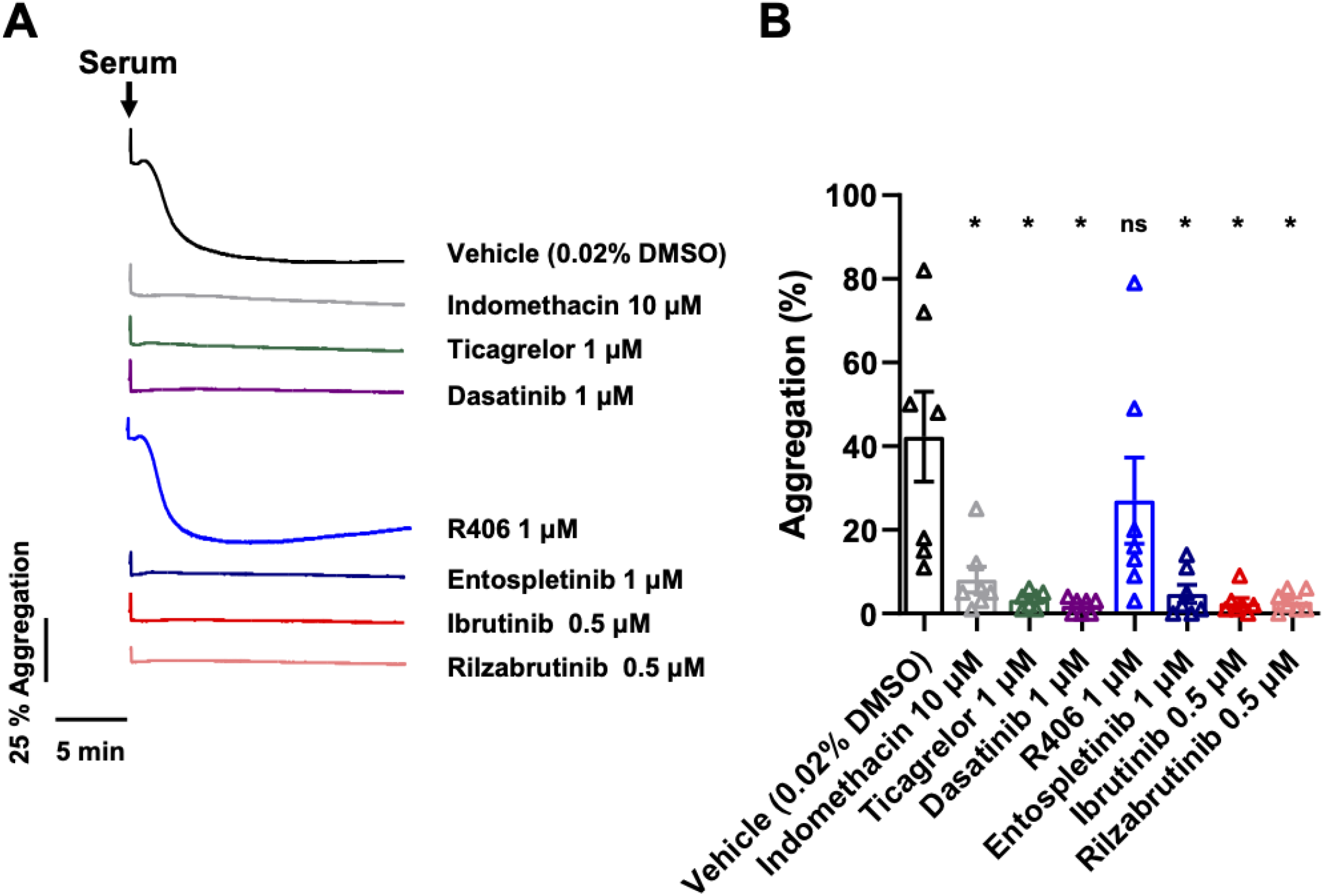
**Inhibition of cyclooxygenase, P**_**2**_**Y**_**12**_, **Src, Syk and Btk block platelet aggregation induced by serum from patients with VITT**. Washed platelets (2×10^8^/ml) were incubated with indomethacin (10 μM), ticagrelor (1 μM), dasatinib (1 μM), R406 (1 μM), entospletinib (1 μM), ibrutinib (0.5 μM), rilzabrutinib (0.5 μM) or vehicle (0.02% DMSO) for 10 minutes then stimulated with serum (1:15, v/v) from patients with VITT. **(A)** Representative aggregation traces and **(B)** quantification of maximum aggregation. Mean ± SEM, n=7. Statistical analysis was by one-way ANOVA with Dunnett multiple comparisons, *p<0.05, ns: non-significant.

## Discussion

In this study we demonstrate that serum from patients with VITT, but not healthy donors, induces platelet aggregation. Platelet aggregation can be abolished by direct blockade of FcγRIIA with IV.3 F(ab) or IVIg, or through inhibition of its downstream signalling kinases. Anti-platelet drugs targeting secondary mediator pathways of platelet activation also prevented aggregation.

All four patients had similar clinical characteristics to those described in the emergent literature for VITT.^5,6^ They all had arterial / venous thrombosis associated with thrombocytopenia, high D-dimers and low fibrinogen. Likewise, all patients were below the age of 50 and presented within two weeks of their first AZD1222 vaccination. Two patients were male and two female. All patients had strongly positive HIT screening tests for anti-PF4 antibodies. Interestingly, in VITT patients when the highly sensitive chemiluminescence-based AcuStar test is used to measure anti-PF4 antibodies it is negative.^17^ This raises the possibility that the anti-PF4 antibodies detected by the ELISA-based Immucor test is a false-positive result.^18^ The Immucor assay is known to produce false positive results in the presence of immune complexes or immunoglobulin aggregates.^19^ This is consistent with our results showing no potentiation of platelet aggregation with the addition of excess PF4.

As previously reported, variable aggregation responses on washed platelets from different healthy donors were observed to sera from patients with VITT.^5^ The low aggregation response to patient 2 serum could potentially be explained by treatment with dexamethasone prior to serum collection. Patients 3 and 4 had not received any treatment prior to sera collection, and elicited much greater responses. As is the case with HIT and other VITT studies, washed platelets from certain healthy donors did not respond.^5,20^ Enhancement of platelet activation to serum from patients with VITT in the presence of PF4 has previously been described by Greinacher et al.^5^, which was not observed in this study. A potential explanation for this difference is the nearly three times higher serum concentrations used by Greinacher et al. Blockade of FcγRIIA preventing platelet aggregation (similar to other reports^5^) implies activation is likely mediated by clustering of the receptor by IgG and immune complexes. We cannot yet however explain the interesting observation that platelet activation was abolished to sera from some patients following heat inactivation. This may reflect a contribution from complement, although normal serum complement levels in VITT patients have been reported.^6^

Immune complexes that activate platelets via FcγRIIA have been reported in critically ill patients with COVID-19.^21^ In these patients, who had been exposed to heparin and displayed thrombocytopenia and thrombosis, HIT was ruled out, due to lack of anti-PF4 antibodies and platelet activation independent of heparin.^21^ Analogous to our findings, platelet activation by these immune complexes could be blocked by both low (0.1 & 0.3 U/ml) and high (100 U/ml) concentrations of heparin.^21^ Current practise is to withhold heparin anticoagulation from patients with VITT.^10^ Our observation of *in vitro* platelet aggregation blockade in the presence of heparin may imply this is not necessary. Anti-SARS-CoV-2 spike protein IgG antibodies from patients with severe COVID-19 have also been shown to induce apoptosis and increase phosphatidyl serine externalisation in platelets mediated by FcγRIIA, although IgG aggregates or immune complexes were not able to be isolated from patient sera.^22^ It is possible that a similar mechanism is occurring in patients with VITT. Activation of FcγRIIA could give rise to phosphatidyl serine exposure and procoagulant platelets which may lead to the extensive thrombosis and thrombocytopenia observed in VITT patients.

We screened a variety of clinically used anti-platelet drugs and inhibitors of kinases downstream of FcγRIIA. Blockade was observed to COX (indomethacin), P2Y_12_ (ticagrelor), Src (dasatinib), Syk (entospletinib) and Btk (ibrutinib and rilzabrutinib) inhibition. These *in vitro* findings, suggest a possible role for prophylactic treatment with aspirin (COX inhibitor) following vaccination. However, with the rarity of the syndrome, the risk of such widespread aspirin usage may cause harms - due to the increased risk of bleeding - that outweigh any theoretical benefit.^17^ Aspirin or ticagrelor use is not currently recommended in patients with confirmed VITT due to the already increased bleeding risk associated with the initial thrombocytopenia, reduced fibrinogen and secondary bleeding from CVST.^23^ If, however, ongoing treatment for VITT is required due to ongoing platelet activation following IVIg treatment once there has been partial restoration of the platelet count, then these anti-platelet agents may have a use at this point.

IVIg treatment has been shown to prevent platelet activation in patients with VITT, but is a scarce and expensive resource.^24^ In patients with confirmed VITT, treatment with the Btk inhibitor rilzabrutinib may be a viable alternative. Rilzabrutinib is currently undergoing phase III clinical trials as a treatment to restore platelet counts in immune thrombocytopenia (ITP) with no reports of bleeding or thrombotic events.^25^ Fostamatinib has been used clinically to treat ITP without causing bleeding (and lowering rates of thrombosis^26^). However, it did not show effectiveness at blocking platelet activation to patient sera *in vitro*. Entospletinib, although not associated with bleeding, is not yet in routine use outside of clinical trials and has not been specifically used in thrombocytopenic patients.^27^ All other inhibitors tested have been associated with an increased risk of bleeding and so their use in thrombocytopenic patients cannot be recommended.^28–30^ Clinical validation of these *in vitro* findings still needs to be performed.

Limitations of this study are the small sample size, and the difference in treatment patients received prior to sample collection. Additionally, due to the limited amount of sera available and the inter-donor variability of platelet responsiveness to serum, only a limited number of conditions could be tested.

In conclusion, serum from patients with VITT activates platelets via FcγRIIA, which can be blocked *in vitro* by anti-platelet therapies and downstream kinase inhibitors. This suggests that prophylaxis in at risk groups with commonly used anti-platelet agents such as aspirin may potentially be able to reduce the impact of VITT and that tyrosine kinase inhibitors may be a treatment for clinically manifest VITT in addition to the current management intervention with corticosteroids and IVIg. These strategies, however, require further study.

## Data Availability

Individual participant data will not be made available.

## Acknowledgements

This work was supported by an Accelerator Grant (AA/18/2/34218) from the British Heart Foundation (BHF). CK is supported by the European Union’s Horizon 2020 Research and Innovation Programme under the Marie Skłodowska-Curie Actions Individual Fellowship grant agreement [No 893262], project PAELLA. SPW holds a BHF Chair (CH03/003). The authors would like to thank Dr Mav Manji (University Hospitals Birmingham NHS Foundation Trust) for help with patient recruitment, Charlotte Stoneley for help sourcing patient blood samples, Matt Roberts for information on the Immucor assay and Principia Biopharma for rilzabrutinib.

## Authorship

CWS and PLRN designed and performed experiments, analysed data and wrote and revised the manuscript. CK designed and performed experiments and revised the manuscript. YD generated reagents and revised the manuscript. SPW revised the manuscript and designed experiments. GCL and WAL recruited patients, revised the manuscript and contributed intellectually. Underlying data was verified by CWS and PLRN.

## References

1 Polack FP, Thomas SJ, Kitchin N, et al. Safety and Efficacy of the BNT162b2 mRNA Covid-19 Vaccine. N Engl J Med 2020; 383: 2603–15.

2 Baden LR, El Sahly HM, Essink B, et al. Efficacy and Safety of the mRNA-1273 SARS-CoV-2 Vaccine. N Engl J Med 2020; 384: 403–16.

3 Voysey M, Clemens SAC, Madhi SA, et al. Safety and efficacy of the ChAdOx1 nCoV-19 vaccine (AZD1222) against SARS-CoV-2: an interim analysis of four randomised controlled trials in Brazil, South Africa, and the UK. Lancet 2021; 397: 99–111.

4 European Centre for Disease Prevention and Control. Total vaccines doses distributed to EU/EAA countries by vaccine product as of 2021-04-08. Eur. Cent. Dis. Prev. Control COVID-19 Vaccine Tracker. https://vaccinetracker.ecdc.europa.eu/public/extensions/COVID-19/vaccine-tracker.html#distribution-tab (accessed April 8, 2021).

5 Greinacher A, Thiele T, Warkentin TE, Weisser K, Kyrle PA, Eichinger S. Thrombotic Thrombocytopenia after ChAdOx1 nCov-19 Vaccination. N Engl J Med 2021; published online April 9. DOI:10.1056/NEJMoa2104840.

6 Schultz NH, Sørvoll IH, Michelsen AE, et al. Thrombosis and Thrombocytopenia after ChAdOx1 nCoV-19 Vaccination. N Engl J Med 2021; published online April 9. DOI:10.1056/NEJMoa2104882.

7 Medicines and Healthcare products Regulatory Agency UK. MHRA issues new advice, concluding a possible link between COVID-19 Vaccine AstraZeneca and extremely rare, unlikely to occur blood clots. Press Release 7/4/2021. 2021. https://www.gov.uk/government/news/mhra-issues-new-advice-concluding-a-possible-link-between-covid-19-vaccine-astrazeneca-and-extremely-rare-unlikely-to-occur-blood-clots (accessed April 7, 2021).

8 Greinacher A, Selleng K, Warkentin TE. Autoimmune heparin-induced thrombocytopenia. J Thromb Haemost 2017; 15: 2099–114.

9 9 Nicolson PLR, Nock SH, Hinds J, et al. Low-dose Btk inhibitors selectively block platelet activation by CLEC-2. Haematologica 2021; 106: 208–19.

10 British Society for Haematology. Guidance produced from the Expert Haematology Panel (EHP) focussed on Covid-19 Vaccine induced Thrombosis and Thrombocytopenia (VITT). 2021. https://b-s-h.org.uk/media/19530/guidance-version-13-on-mngmt-of-thrombosis-with-thrombocytopenia-occurring-after-c-19-vaccine_20210407.pdf (accessed April 7, 2021).

11 Warkentin TE. High-dose intravenous immunoglobulin for the treatment and prevention of heparin-induced thrombocytopenia: a review. Expert Rev Hematol 2019; 12: 685–98.

12 Rubino JG, Arnold DM, Warkentin TE, Smith JW, Kelton JG, Nazy I. A comparative study of platelet factor 4-enhanced platelet activation assays for the diagnosis of heparin-induced thrombocytopenia. J Thromb Haemost 2021; 19: 1096–102.

13 Vayne C, Guery E-A, Kizlik-Masson C, et al. Beneficial effect of exogenous platelet factor 4 for detecting pathogenic heparin-induced thrombocytopenia antibodies. Br J Haematol 2017; 179: 811–9.

14 Padmanabhan A, Jones CG, Curtis BR, et al. A Novel PF4-Dependent Platelet Activation Assay Identifies Patients Likely to Have Heparin-Induced Thrombocytopenia/Thrombosis. Chest 2016; 150: 506–15.

15 Warkentin TE, Arnold DM, Nazi I, Kelton JG. The platelet serotonin-release assay. Am J Hematol 2015; 90: 564–72.

16 Arman M, Krauel K. Human platelet IgG Fc receptor FcγRIIA in immunity and thrombosis. J Thromb Haemost 2015; 13: 893–908.

17 Scully M, Singh D, Lown R, et al. Pathologic Antibodies to Platelet Factor 4 after ChAdOx1 nCoV-19 Vaccination. N Engl J Med 2021; published online April 16. DOI:10.1056/NEJMoa2105385.

18 Favaloro EJ, McCaughan G, Mohammed S, et al. HIT or miss? A comprehensive contemporary investigation of laboratory tests for heparin induced thrombocytopenia. Pathology 2018; 50: 426–36.

19 Immucor. PF4 IgG assay [package insert]. USA: Immucor; 2017. 2017.

20 Warkentin TE, Hayward CPM, Smith CA, Kelly PM, Kelton JG. Determinants of donor platelet variability when testing for heparin-induced thrombocytopenia. J Lab Clin Med 1992; 120: 371–9.

21 Nazy I, Jevtic SD, Moore JC, et al. Platelet-activating immune complexes identified in critically ill COVID-19 patients suspected of heparin-induced thrombocytopenia. J Thromb Haemost 2021; : 1–6.

22 Althaus K, Marini I, Zlamal J, et al. Antibody-induced procoagulant platelets in severe COVID-19 infection. Blood 2021; 137: 1061–71.

23 Zheng SL, Roddick AJ. Association of Aspirin Use for Primary Prevention With Cardiovascular Events and Bleeding Events: A Systematic Review and Meta-analysis. JAMA 2019; 321: 277–87.

24 Misbah SA, Murphy MF, Pavord S, et al. Outcome of national oversight of intravenous immunoglobulin prescribing in immune thrombocytopenia. J Clin Pathol 2020; 0: 2019–20.

25 Kuter DJ, Boccia R V, Lee E-J, et al. Phase I/II, Open-Label, Adaptive Study of Oral Bruton Tyrosine Kinase Inhibitor PRN1008 in Patients with Relapsed/Refractory Primary or Secondary Immune Thrombocytopenia. Blood 2019; 134: 87.

26 Cooper N, Altomare I, Kreychman Y, et al. Abstracts. PB1356 |Immune Thrombocytopenia Treatment with Fostamatinib, a Spleen Tyrosine Kinase Inhibitor: Reducing the Risk of Thrombosis. Res Pract Thromb Haemost 2020; 4: 1–1311.

27 Awan FT, Thirman MJ, Patel-Donnelly D, et al. Entospletinib monotherapy in patients with relapsed or refractory chronic lymphocytic leukemia previously treated with B-cell receptor inhibitors: results of a phase 2 study. Leuk Lymphoma 2019; 60: 1972–7.

28 Becker RC, Bassand JP, Budaj A, et al. Bleeding complications with the P2Y12 receptor antagonists clopidogrel and ticagrelor in the PLATelet inhibition and patient Outcomes (PLATO) trial. Eur Heart J 2011; 32: 2933–44.

29 Shatzel JJ, Olson SR, Tao DL, McCarty OJT, Danilov A V., DeLoughery TG. Ibrutinib-associated bleeding: pathogenesis, management and risk reduction strategies. J Thromb Haemost 2017; 15: 835–47.

30 Quintás-Cardama A, Kantarjian H, Ravandi F, et al. Bleeding diathesis in patients with chronic myelogenous leukemia receiving dasatinib therapy. Cancer 2009; 115: 2482–90.

